# Investigating AI’s potential in risk-stratifying lung cancer screenings for extending screening intervals and reducing workload: A retrospective analysis of a tertiary hospital cohort

**DOI:** 10.64898/2026.11.03.26367146

**Authors:** Ali Nowroozi, Masha Bondarenko, Sukhmanjit S Brar, Brian Haas, James Brown, Alison Rustagi, Brian Graham, Mollie Hudson, George Su, Thienkhai H. Vu, Adam Yala, Jae Ho Sohn

## Abstract

**Objective:** We aimed to 1) evaluate whether AI algorithms could identify a very low-risk sub-population Lung-RADS 1/2 cases suitable for a safe switch to biennial screening, and 2) externally validate Sybil, an imaging-based deep learning lung cancer risk prediction tool.

**Materials and methods:** We gathered all lung cancer screenings (LCSs) from a tertiary safety-net hospital (n=3120, up to September 2024), manually extracted relevant clinical data, and analyzed Sybil’s performance across various subgroups, with a specific focus on Lung-RADS 1 and 2 cases. We simulated LCS interval extension using Sybil and the Brock model risk scores calibrated on a subset of the National Lung Screening Trial (NLST). Confidence intervals were obtained through bootstrapping.

**Results:** The median follow-up was 2.3 years (interquartile range 1.5–3.1), with a total 6-year cancer incidence rate of 2.3% (72/3120). Lung-RADS 1 and 2 cases comprised 90.5% of our scans. Sybil’s overall area under the curve (AUC) ranged from 0.819 to 0.912; while its 2-year AUC for Lung-RADS 1 and 2 cases was 0.746. In the screening interval extension simulation, calibrating for a 30% shift to biennial screening translated into 33.8% of negative screens in our data being safely extended using Sybil, with no delayed cancers. The Brock model demonstrated an AUC of 0.513 on our data, with a sensitivity of 18.2% using the 1.5% threshold.

**Conclusion:** Sybil has the potential to safely extend LCS intervals for low-risk individuals, reducing radiology workload and opening spots for life-saving new screenings.

**Key points:**

- We investigated whether AI risk stratification tools could identify the lowest-risk individuals undergoing lung cancer screening suitable for switching to a biennial screening schedule.
- In our simulations, Sybil reduced follow-up screening load by at least 30% through switching the lowest-risk Lung-RADS 1 and 2 cases to biennial screening without delayed cancer diagnoses.
- Very low-risk individuals, as predicted by AI algorithms, could potentially be placed on biennial screening schedules, reducing radiology department workload and freeing time for new patients.

## Introduction

The United States Preventive Services Task Force (USPSTF) has recommended lung cancer screening (LCS) since 2013 based on the results of the National Lung Screening Trial (NLST) and other randomized controlled screening trials (1). In recent years, with ever-rising imaging volumes, radiology practices have been facing an unprecedented workforce shortage (2). The rollout of lung cancer screening has also contributed to the workforce-volume mismatch, as screening contributes directly to increased imaging volume (3) as well as resultant downstream work of clinical consultations, clinical decision-making conferences, follow-up scans, and lung biopsies. These challenges are often exacerbated in lower-resource settings, such as safety-net hospitals. Previous studies have demonstrated the utility of artificial intelligence (AI) in reducing lung cancer screening workload by serving as the first reader (4,5).

The USPSTF guidelines do not provide recommendations on how screenings should be followed; however, they mention Lung-RADS as an option. In practice, the Lung-RADS criteria are widely adopted as the management framework. According to the criteria, lowest-risk scans (i.e., Lung-RADS 1 and 2) are assigned a 1-year interval, while higher-risk individuals are assigned 6-month or 3-month intervals or a direct biopsy recommendation. Unlike the most recent version of Lung-RADS, the original version was not associated with a companion paper (6–9), and studies continue to optimize the recommendations.

Some studies and guidelines have suggested that individuals at lower risk of developing lung cancer, who comprise the majority of LCS cases, may be suitable for extended screening intervals without delaying cancer diagnoses (10–12). However, there are currently no guideline-endorsed risk-stratification tools for identifying this subgroup, and assigning individualized screening intervals is a topic of active investigation (13). Personalized breast cancer screening schedules using an AI risk prediction algorithm (Mirai) have previously been investigated (14,15). Furthermore, while the 1-year performance of Lung-RADS is well documented (16), very few studies have reported the multi-year risk of malignancy across Lung-RADS categories, especially in the US (17), despite its crucial impact on follow-up schedules and policies.

In this study, we pursued 2 main goals: 1) investigating through simulation whether two lung cancer risk prediction tools (deep-learning and imaging-based Sybil (18) and the regression-based Brock model (19)) could identify Lung-RADS 1 and 2 cases at sufficiently low risk to extend these patients’ screening interval to 2 years, and 2) externally validating Sybil on our data.

## Methods

### Lung cancer risk prediction

Sybil (18) is an image-based deep learning lung cancer risk prediction model that provides 1-to 6-year risk scores (i.e., risk of developing cancer within that period of time after a screening instance) ranging from 0 to 1 for screening scans and is accessible via https://github.com/reginabarzilaygroup/Sybil. We chose the Brock model (19,20), which predicts a nodule’s risk score using demographics and radiological features, as a secondary risk-prediction tool and a comparison reference. The Brock model is being used in the International Lung Screening Trial (ILST) to designate screening candidates to biennial screening (21).

### Screening interval extension simulation

We simulated the feasibility and safety of assigning very low risk individuals to biennial screening by further risk-stratifying Lung-RADS 1 and 2 cases, who already have low risk of cancer, using our AI tools. For this task, we used the 2-year Sybil risk score, as we intend to identify the risk of developing cancer within 2 years. We use the 1.5% threshold for the Brock model, which is the threshold being used in the ILST. The main outcome of interest was the trade-off between reduced workload (i.e., fewer scans and biopsies) and negative health outcomes (i.e., delayed cancers due to not screening at the 1-year mark). We defined at-risk-of-delay cancers as cancers that could be detected if the patient received the normal 1-year interval screening. The definition was not the temporal 2^nd^-year cancer cases (i.e., between 365 and 730 days from the scan); they were determined through two criteria derived from the data to increase precision: 1) If the subject received an appropriate follow-up screen (9 – 18 months after the initial screen to allow for variations) and the patient was diagnosed with cancer within 3 months of that screen or the scan demonstrated the cancerous lesion, or 2) if the subject did not have an appropriate follow-up screening but was diagnosed with cancer between 1 and 2 years after the initial scan (we assumed that these cancers would have been detectable). Delayed cancers were defined as at-risk-of-delay cases assigned to biennial screening. Since Sybil provides continuous scores, we applied cutoffs based on the NLST data as our calibration dataset. We did this within two frameworks, following a similar methodology in a previous study (22): the cutoff was set based on targeting 1) maximum allowed delayed cancer rate (i.e., 1 - sensitivity), or 2) minimum rate of reduction in follow-up scans of non-interval-cancer cases (i.e., specificity). We also investigated whether any negative interval biopsies (defined similarly to delayed cancers) could have been avoided as a means of reducing workload.

For delayed cancers and averted biopsies, due to their low denominators, we also report a model-based probabilities based on the Sybil scores and the selected thresholds, in addition to the raw observed rates. Further description can be found in the statistical analysis section.

### Data

#### Our institution

This is a HIPAA-compliant IRB-approved (IRB approval 17-22317) retrospective cohort study that follows the STROBE checklist. We retrieved all LCS CT studies from the Zuckerberg San Francisco Hospital’s radiological database up to September 2024 (to allow at least 1 year of follow-up) (supplementary methods). The only exclusion criterion was restricted access to the patient’s electronic health records. Reports with no explicit Lung-RADS assignment (7%) were assigned a score based on the report description and according to Lung-RADS v2022 by a medical doctor with 2 years of radiology research postdoctoral training, under the supervision of a fellowship-trained cardiothoracic radiologist. Age and sex were extracted as demographic data, and last available patient encounter with any provider, lung biopsy, and lung cancer diagnosis dates (if applicable) were manually extracted from patient charts to define clinical outcomes. Follow-up duration was defined as the interval between the date of the scan and the last observed encounter. Family history of lung cancer based on the electronic health records and emphysema and nodules details based on the radiology report were also extracted for a subset of Lung-RADS 2 cases (all positive second year cancer cases + random 100 cases with no cancer within 2 years given the difficulty in collecting the data needed for this model such as family history and radiological features from the reports) to calculate the Brock (19,20) risk scores. Lung-RADS 1 cases were excluded from the Brock analysis, as nodules with benign features were excluded from the Brock study and were automatically assigned to biennial screening in the ILST (19,21).

#### NLST

We used a subset of Lung-RADS-assigned NLST cases, which were also used in testing Sybil (18), as our calibration dataset in the interval extension simulation, described below. These cases were Lung-RADS 1 and 2 cases that were not diagnosed with cancer in the first year, and had at least 2 years of follow-up or were diagnosed with cancer in the 2^nd^ year. This yielded 5542 screenings from 2139 subjects, forming our calibration dataset. More details on calibration can be found in the sections below.

### Retrospective external validation of Sybil

We validated Sybil scores on our institution’s data and in various subgroups. In all analyses, each score was used only with data for the years corresponding to that score; for instance, the 3-year score was only validated on cancer outcomes within the first 3 years after a scan, and only on cases with follow-ups up to that point.

### Statistical analysis

All main analyses were conducted for screening instances, not persons (i.e., two screening sessions from the same person count as separate cases), as Sybil provides risk scores for scans, and screening interval decisions are also made based on individual scans. Confidence intervals for proportions were calculated using Wilson score interval with continuity correction. Performance measures for predicting binary outcomes included accuracy, sensitivity, and specificity. Discriminatory performances of Sybil’s continuous scores were assessed using the area under the curve (AUC) metric. Confidence intervals for the AUCs were calculated via 1000-resample bootstrapping. To model the delayed cancers and averted biopsies, we assumed a normal distribution for the logit-transformed 2-year Sybil scores of all the at-risk-of-delay and biopsied cases and the modeled rates were the probability of a risk score < the threshold. The 95% CI was derived by 2000 resamples of the NLST calibration data for the cutoff and, for each cutoff, 20 resamples with replacement of our institution’s screenings, with the mean and standard deviation of the logit-transformed scores re-estimated in each resample. The 2.5th and 97.5th percentiles of the resulting 40,000 estimates formed the interval. This accounts for uncertainty in both the cutoff and the score distribution. Analyses were performed in Python 3.12.10 using *scikit-learn* (v1.8.0), *statsmodels* (v0.14.6), and scipy (v1.17.0) libraries.

## Results

### Overall descriptives

The final dataset comprised 3120 screenings (from 2125 patients) dating back to March 2015, 38 of which Sybil failed to provide a score for (Figure 1). The mean age of cases was 63.1 ± 6.9 years, of which 2625 (84.1%) were male. Total follow-up was 7785.3 years with a median of 2.3 years (interquartile range (IQR) = 1.5 – 3.1 years). Within 6 years, 72 cases (2.3%) were diagnosed with lung cancer. Overall, Lung-RADS 0 to 4x cases comprised 1.4%, 22.0%, 68.5%, 3.8%, 2.5%, 1.7%, and 0.3% of the scans, respectively (Figure 2). The share of Lung-RADS 1, 2, 3, and 4a cases that received their next screening in a timely manner (within 2 years, 2 years, 1 year, and 6 months, respectively, to allow for variations) was 38.7%, 48.9%, 54.8%, and 40.8%, respectively. Visualization of intervals to the next screening is shown in Figure 3. Table 1 and Figure 4 show cancer statistics, classified by Lung-RADS, for all scans. One-year cancer incidence after only baseline scans was 1.8% (35/1927), and there were 14 and 11 cancer cases in the first 2 years after all and baseline-only Lung-RADS 1 and 2 scans (Table 1, supplementary table S1).

**Figure 1.**
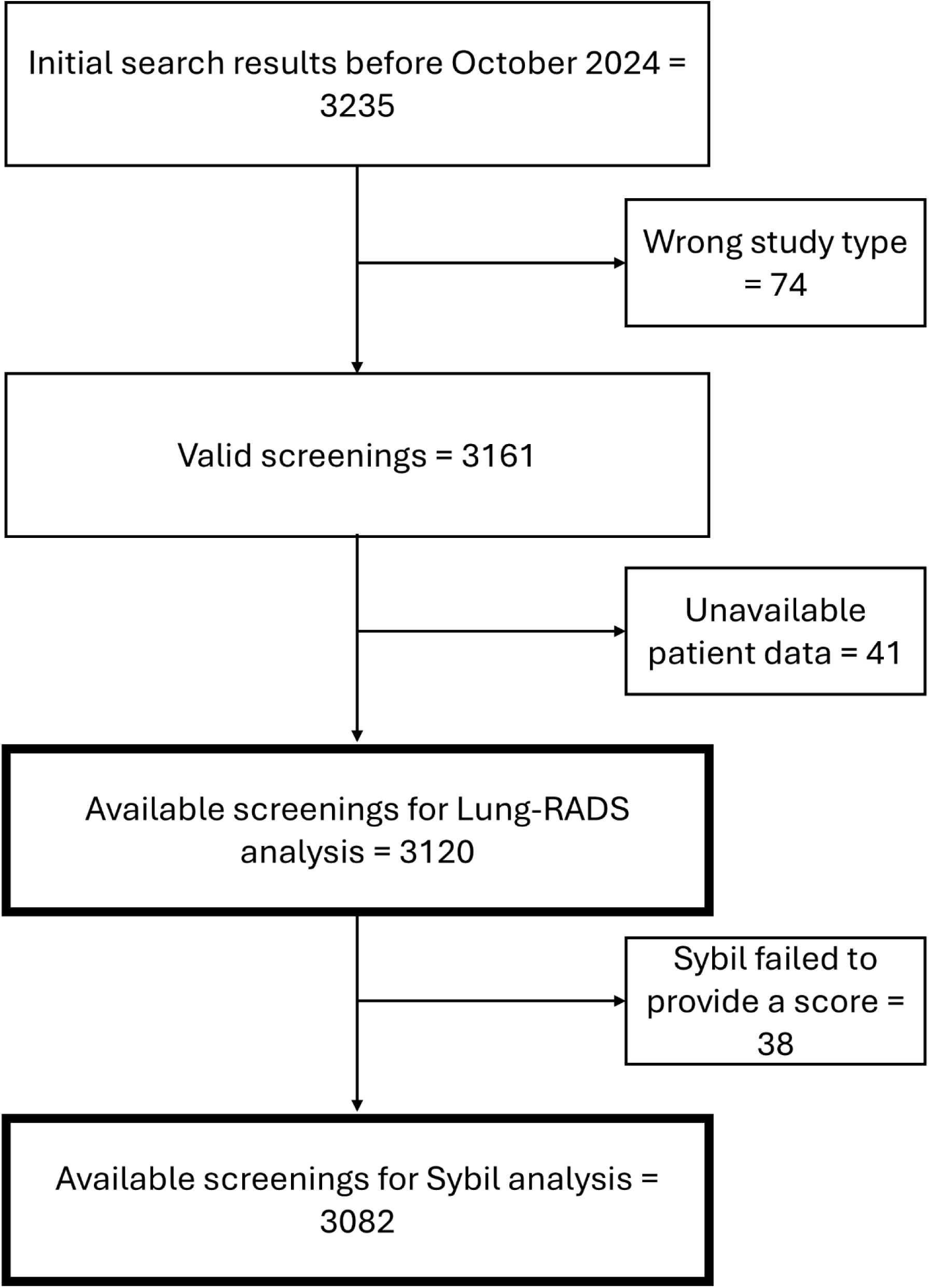
Included cases flow diagram.

**Figure 2.**
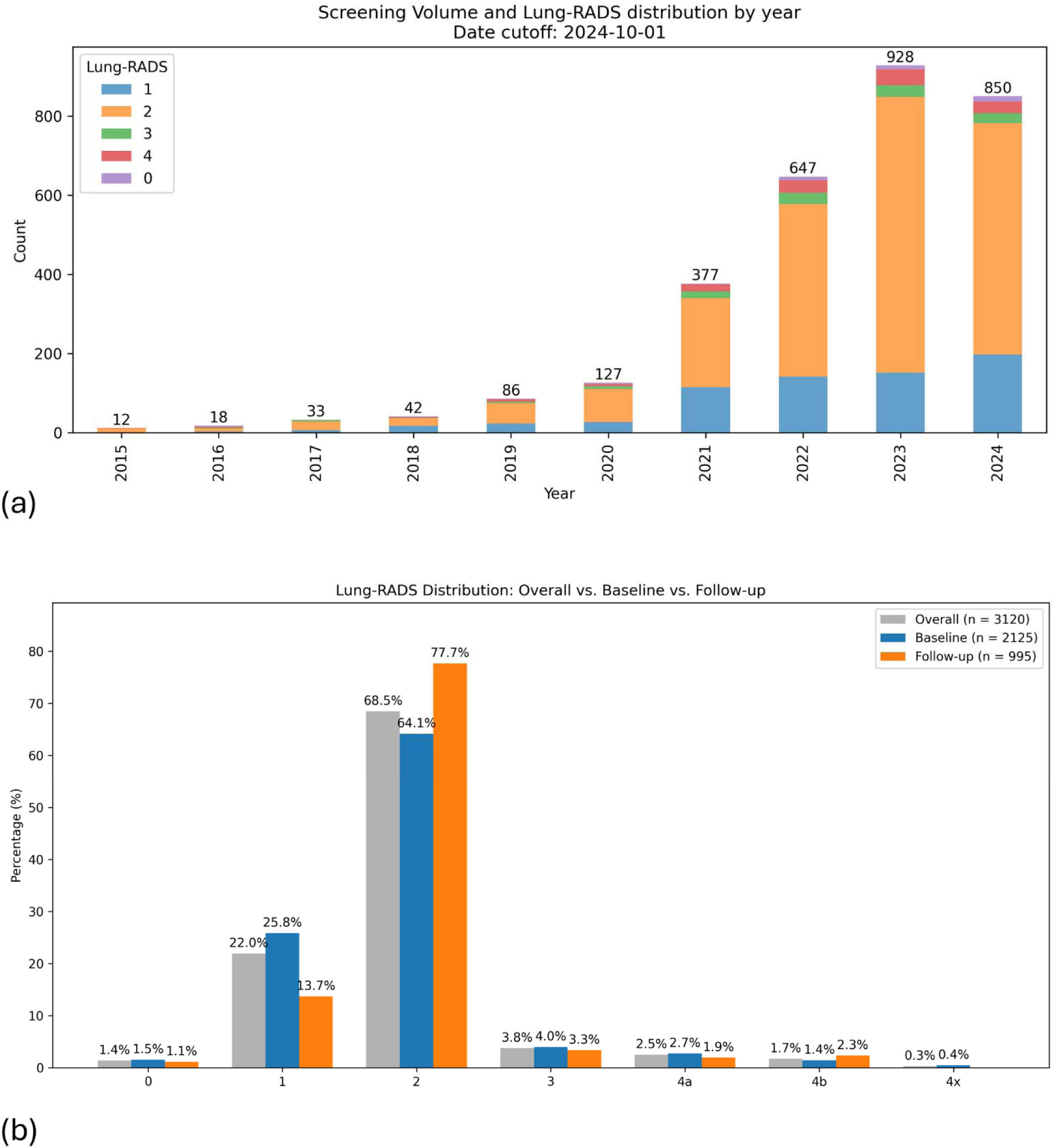
Screening cohort descriptive statistics: a) Screening volume and Lung-RADS distribution by year, b) Lung-RADS distribution by scan instance category: overall, baseline, and follow-up.

**Figure 3.**
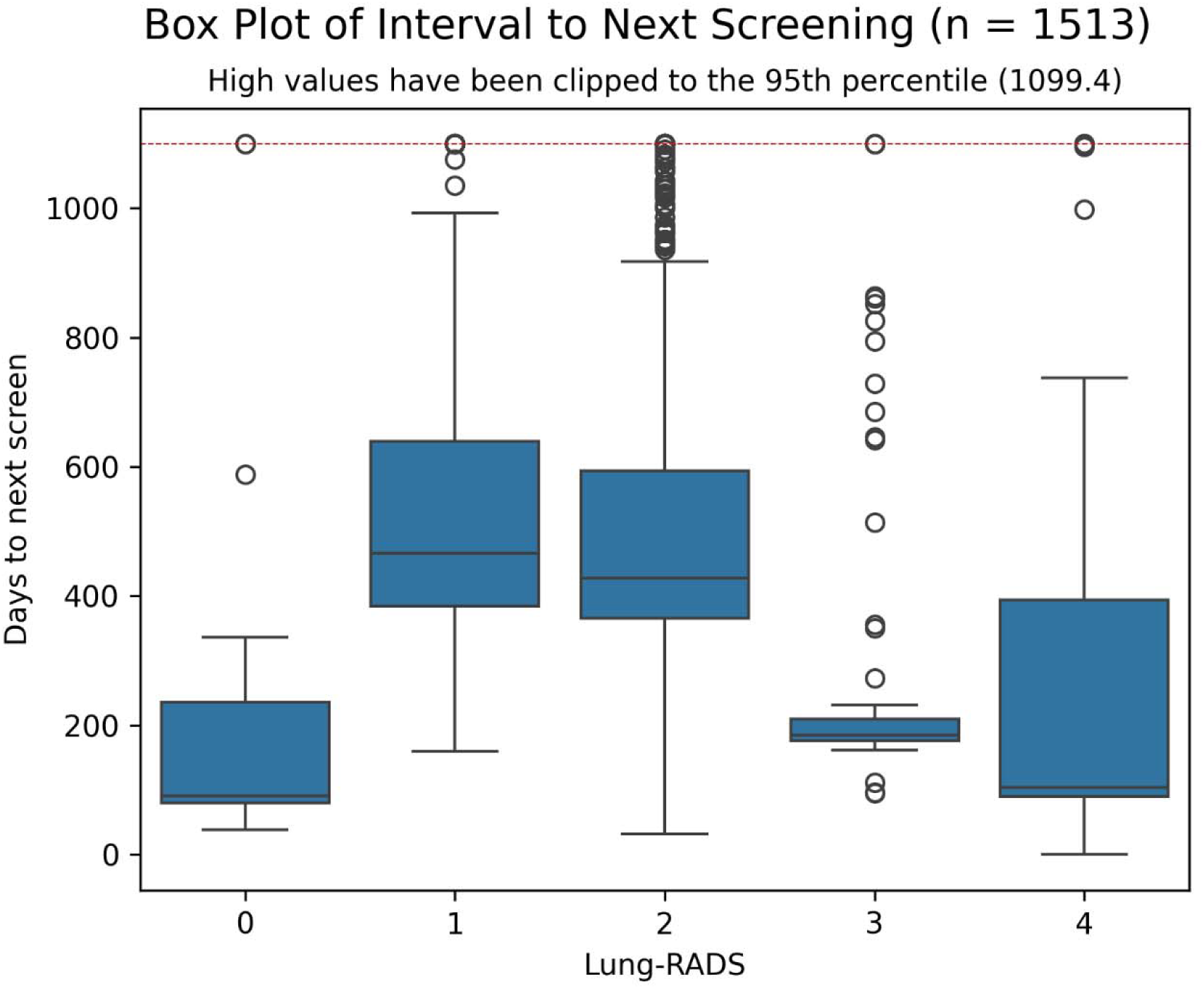
Box plot of interval to next screening. The values have been clipped to the 95^th^ percentile (1099.4, indicated by the red line).

**Figure 4.**
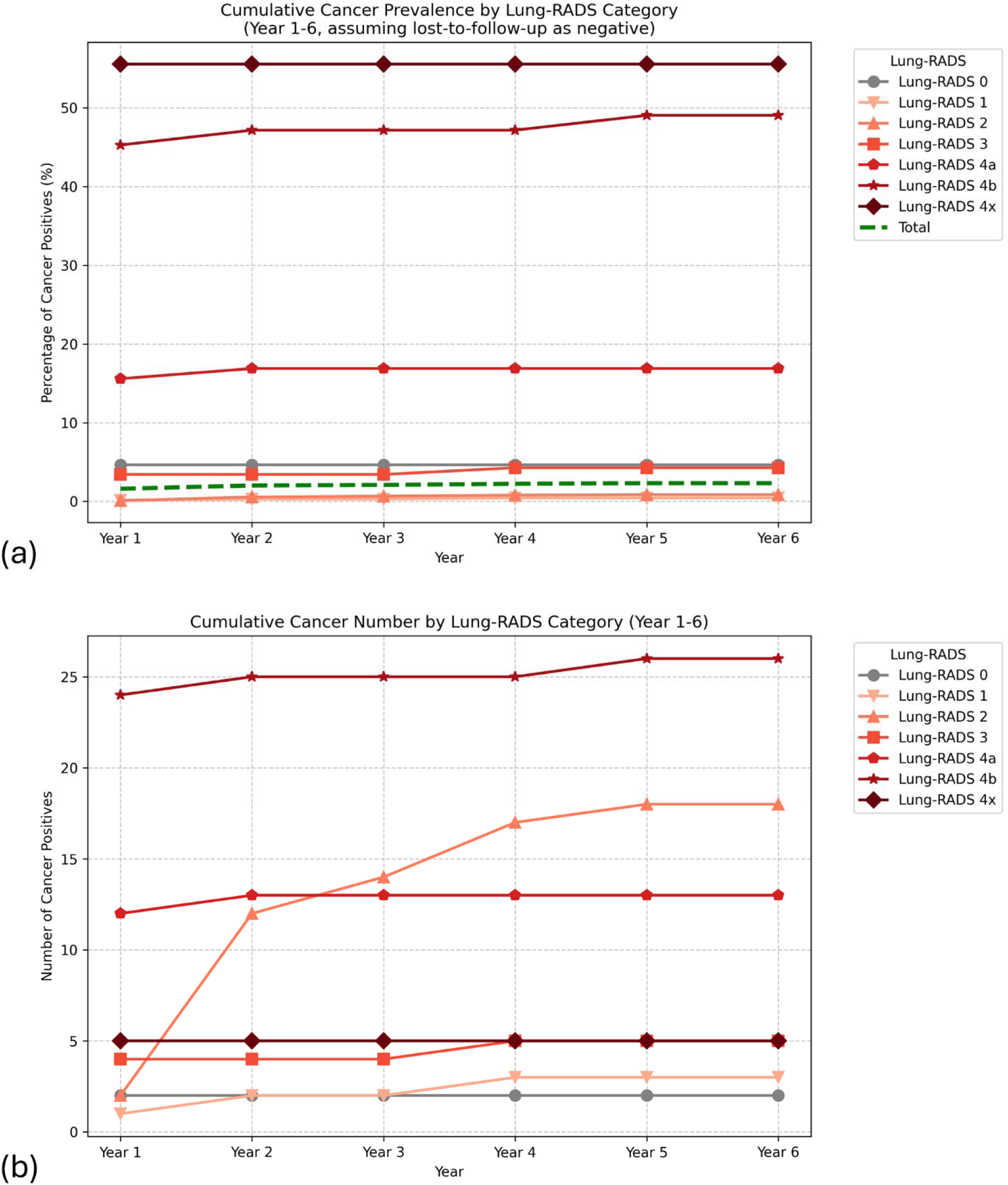
The 6-year trend of lung cancer statistics by Lung-RADS: a) Cumulative cancer prevalence (%); lost to follow-up cases were assumed negative); green line indicates all Lung-RADS categories combined, b) Cumulative cancer absolute number.

**Table 1.** Annual lung cancer incidence. Data for each year includes only patients with follow-up through that year, except for the final total column, which assumes all lost to follow-up cases as negative. Years are chronological and do not represent screening-based years, as participants have varying screening schedules.

| **Lung-RADS** | **New lung cancer diagnoses (incidence %, 95% CI)** |  |  |  |  |  |  |
|---|---|---|---|---|---|---|---|
|  | **Year 1** | **Year 2** | **Year 3** | **Year 4** | **Year 5** | **Year 6** | **Total (Assuming lost to follow-up as negative)** |
| **0** | 2/34 (5.9%, 1.0-21.1) | 0/16 (0.0%, 0.0-24.1) | 0/12 (0.0%, 0.0-30.1) | 0/8 (0.0%, 0.0-40.2) | 0/4 (0.0%, 0.0-60.4) | 0/3 (0.0%, 0.0-69.0) | 2/43 (4.7%, 0.8-17.1) |
| **1** | 1/620 (0.2%, 0.0-1.0) | 1/465 (0.2%, 0.0-1.4) | 0/223 (0.0%, 0.0-2.1) | 1/108 (0.9%, 0.0-5.8) | 0/45 (0.0%, 0.0-9.8) | 0/25 (0.0%, 0.0-16.6) | 3/685 (0.4%, 0.1-1.4) |
| **2** | 2/1908 (0.1%, 0.0-0.4) | 10/1387 (0.7%, 0.4-1.4) | 2/516 (0.4%, 0.1-1.6) | 3/230 (1.3%, 0.3-4.1) | 1/106 (0.9%, 0.0-5.9) | 0/57 (0.0%, 0.0-7.9) | 18/2136 (0.8%, 0.5-1.4) |
| **3** | 4/103 (3.9%, 1.3-10.2) | 0/62 (0.0%, 0.0-7.3) | 0/38 (0.0%, 0.0-11.4) | 1/18 (5.6%, 0.3-29.4) | 0/8 (0.0%, 0.0-40.2) | 0/5 (0.0%, 0.0-53.7) | 5/117 (4.3%, 1.6-10.2) |
| **4a** | 12/67 (17.9%, 10.0-29.6) | 1/37 (2.7%, 0.1-15.8) | 0/22 (0.0%, 0.0-18.5) | 0/10 (0.0%, 0.0-34.5) | 0/7 (0.0%, 0.0-43.9) | 0/4 (0.0%, 0.0-60.4) | 13/77 (16.9%, 9.6-27.5) |
| **4b** | 24/51 (47.1%, 33.2-61.4) | 1/17 (5.9%, 0.3-30.8) | 0/7 (0.0%, 0.0-43.9) | 0/2 (0.0%, 0.0-80.2) | 1/2 (50.0%, 2.7-97.3) | 0 (n/a) | 26/53 (49.1%, 35.3-63.0) |
| **4x** | 5/5 (100.0%, 46.3-100.0) | 0 (n/a) | 0 (n/a) | 0 (n/a) | 0 (n/a) | 0 (n/a) | 5/9 (55.6%, 22.7-84.7) |
| **Total** | 50/2788 (1.8%, 1.3-2.4) | 13/1984 (0.7%, 0.4-1.1) | 2/818 (0.2%, 0.0-1.0) | 5/376 (1.3%, 0.5-3.3) | 2/172 (1.2%, 0.2-4.6) | 0/94 (0.0%, 0.0-4.9) | 72/3120 (2.3%, 1.8-2.9) |

### Screening interval simulation extension

We simulated transitioning the lowest-risk Lung-RADS 1 and 2 cases to a 2-year screening interval by determining cut-offs for Sybil scores based on the NLST dataset (i.e., the calibration dataset). Maximum delayed cancer rates of 0%, 5%, and 10% for predicting 2nd-year cancer in the calibration dataset translated into 0/12 second-year cancers being delayed in testing, with modeled delayed cancer rates of 1.2% (95% CI 0.0 – 5.6%), 2.3% (0 – 12.2%), and 2.3% (0.1 – 26.5%), respectively (Table 2). Targeting workload reductions of 10%, 20%, and 30% resulted in 0 delayed cancers in the test group (modeled 1.2% (0 – 3.9%), 1.2% (0 – 4.8%), and 4.9% (0.3 – 14.2%), respectively), and yielded up to 33.8% (31.7 – 36.0%) reduction of negative screenings and 1/9 (11.1%) averted negative biopsies.

**Table 2.** Sybil simulation results assessing the potential of setting a target maximum delayed detection rate. **(a) and a target minimum interval extension rate (b).** Delayed cancers were defined by: 1) If the subject received an appropriate follow-up screen (9 – 18 months after the initial screen to allow for variations) and the patient was diagnosed with cancer within 3 months, or 2) if the subject did not have an appropriate follow-up screening but was diagnosed with cancer between 1 and 2 years after the initial scan (we assumed that these cancers would have been detectable).

| (a) Calibration dataset target maximum (and actual) allowed delayed cancer detection (100% - sensitivity) | Threshold (95% CI) | Test delayed cancers, % (modeled, 95% CI) | Test specificity (safely extended screenings), % (bootstrap 95% CI) | Averted negative biopsies, % (modeled, 95% CI) |
|---|---|---|---|---|
| 10% (2.6%) | 0.00205 (0.00205 – 0.00679) | 0/12, 0% (2.3%, 0.1 – 26.5%) | 303/1820, 16.6% (15.0 – 58.9%) | 0/9, 0% (6.4%, 0.0 – 27.2%) |
| 5% (2.6%) | 0.00205 (0.00157 – 0.00387) | 0/12, 0% (2.3%, 0 – 12.2%) | 303/1820, 16.6% (5.1 – 36.4%) | 0/9, 0% (6.4%, 0.0 – 17.3%) |
| 0% | 0.00157 (0.00157 – 0.00205) | 0/12, 0% (1.2%, 0 – 5.6%) | 104/1820, 5.7% (4.8 – 18.5%) | 0/9, 0% (4.6%, 0.0 – 13.1%) |
| **(b) Calibration dataset target minimum (and actual) negative screening interval extension rate (specificity)** | **Threshold (95% CI)** | **Test delayed cancers, % (modeled, 95% CI)** | **Test specificity (safely extended screenings), % (bootstrap 95% CI)** | **Averted negative biopsies, % (modeled, 95% CI)** |
| 10% (20.3%) | 0.00159 (0.00159 – 0.00171) | 0/12, 0% (1.2%, 0 – 3.9%) | 299/1820, 16.4% (14.7 – 18.1%) | 0/9 (4.6%, 0.0 – 11.7%) |
| 20% (20.3%) | 0.00159 (0.00159 – 0.00209) | 0/12, 0% (1.2%, 0 – 4.8%) | 299/1820, 16.4% (14.8 – 25.2%) | 0/9 (4.6%, 0.0 – 12.5%) |
| 30% (36.1%) | 0.00287 (0.00287 – 0.00387) | 0/12, 0% (4.9%, 0.3 – 14.2%) | 616/1820, 33.8% (31.7 – 36.0%) | 1/9, 11.1% (9.5%, 0.0 – 21.0%) |
| 40% (41.0%) | 0.00403 (0.00403 – 0.00556) | 1/12, 8.3% (9.5%, 1.0 – 21.7%) | 715/1820, 39.3% (37.1 – 52.0%) | 1/9, 11.1% (13.7%, 0.0 – 27.0%) |

The Brock model showed an AUC of 0.513 (0.333 – 0.721) for predicting 2^nd^-year lung cancers (Supplementary Figure S3). With a threshold of 1.5%, 9 out of 11 (81.8%) cases had a delayed lung cancer diagnosis.

We performed a sensitivity analysis by limiting only to baseline studies, as this is how the Brock model is being used in ILST. In the Sybil analysis, there were 0/9 delayed cancers up to 10% delayed cancer rate allowed, with modeled delays of 1.2% (0 – 18.1%), 1.2 (0 – 19.2%), and 1.2 (0 – 24.1%) for 0%, 5%, and 10%, respectively. Workload reductions of up to 40% did not delay any cancers, with modeled delay rates of 0.6% (0 – 3.1%), 0.6% (0 – 3.1%), 1.3% (0 – 10.0%), and 6.2% (0 – 17.9%), for 10%, 20%, 30%, and 40% workload reductions, respectively. Brock model AUC for baseline cases was 0.521 (0.282 – 0.783), with 6/8 cancers delayed (sensitivity = 25.0%).

### Retrospective Sybil external validation

Sybil’s AUC for predicting cancer ranged from 0.819 (95% CI 0.749 – 0.879) (6-year score) to 0.912 (0.859 – 0.954) (1-year score) (Figure 5). Nonetheless, when limited to Lung-RADS 1 and 2 cases, the performance of Sybil’s 2-year score was modest, with an AUC of 0.746 (0.636 – 0.850). The best performance was observed in the Lung-RADS 3 subcategory (AUC = 0.965 (0.885 – 1.000)).

**Figure 5.**
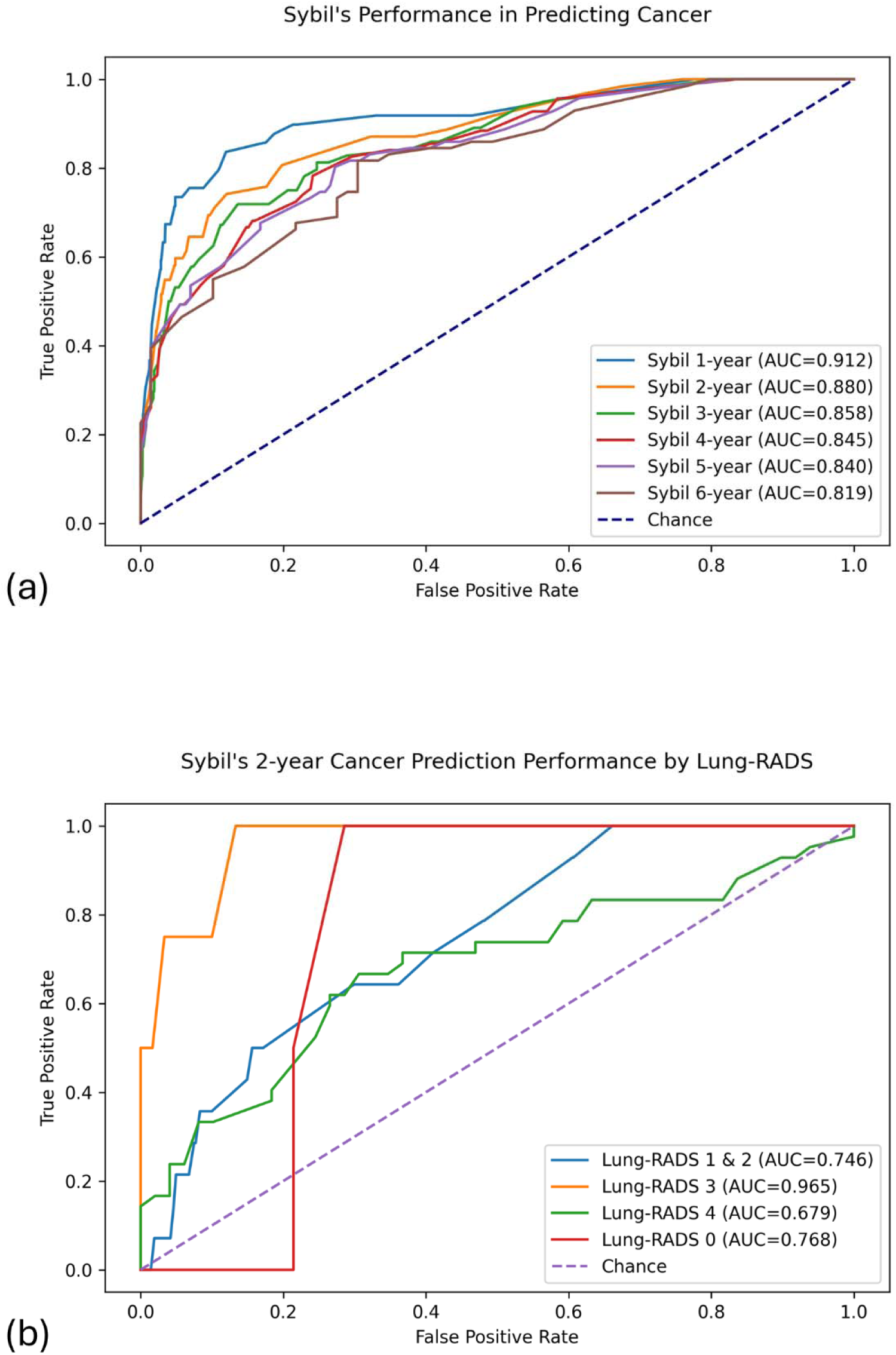
Receiver operating characteristic (ROC) curves for Sybil’s performance: a) All years, b) 2-year prediction by Lung-RADS. Data used for each score matches the year of the score (e.g., 3-year score results were based on data for cases with at least 3 years of follow-up and at the 3-year mark).

## Discussion

In this study, we provided an overview of LCS cases at our tertiary safety-net hospital and evaluated whether AI tools could help identify suitable cases for switching to biennial lung cancer screening. At our institution, lung cancer screening volume increased rapidly in recent years, with more than a 40% increase from 2022 to 2023, and the number of screenings through September 2024 almost matching the 2023 total. This example of the rapidly growing burden on radiology departments emphasizes the need for novel methods, such as AI, to assist in the process.

Simulations showed promise for Sybil in assigning biennial screening intervals for the lowest-risk patients. In our data, up to 30% of screenings could be switched to a biennial schedule without any delayed cancers, with the caveat of a limited sample size of positive cases.

The 1.5% threshold for the Brock model, on the other hand, did not perform well on our data and delayed more than 80% of 2^nd^-year cancer diagnoses. A previous analysis on this model showed that the 1.5% threshold missed 30% of the cancers developed within 2 years (23). Another analysis demonstrated that for 0% delayed cancers, only 2.7% of the cancer-free cases will be assigned to biennial screening (vs 5.7% for Sybil), and for 25.8% of the cancer-free cases to be assigned to biennial screening, 9.8% of the cancer cases will potentially be delayed (vs 0% for our data with 30% safe biennial assignment).

Sybil performed well overall at predicting cancer (AUCs >0.8 across all years and >0.85 for years 1-3), particularly given that our case was an external validation scenario. While its discriminatory performance for our main subgroup of interest (i.e., Lung-RADS 1 and 2 cases) was not satisfactory in terms of AUC, the receiver operating characteristic (ROC) curve showed a range of very high sensitivity with fair specificity, resulting in the favorable simulation results. This finding highlights the importance of evaluating beyond the raw AUC values and focus on performance at thresholds relevant to the clinical question.

The large number of benign lung cancer screenings makes identifying and extending the interval for even a small portion of them highly valuable for reducing workload and radiation. Hence, it is logical to consider a high-sensitivity, low-specificity threshold for risk-stratification scores. In our Sybil simulations, we reduced second-year screenings by 30% without delaying cancer diagnoses. Assuming 1000 annual lung cancer screenings, the 30% reduction rate frees up 270 screening spots (0.3 * 1000 (total screenings) * 0.9 (share of Lung-RADS 1 and 2)) that could be used for new baseline screenings. The NELSON and MILD trial results indicated a number needed to scan of 130 and 167 to avert one lung cancer death, respectively (24,25), meaning that approximately 2 lung cancer deaths would be averted by if 270 new patients fill the spots and adhere to their screenings.

We used Sybil in our study, which is an open-source model. A few previous studies have utilized proprietary deep learning models to extend lung cancer screening intervals. Landy et al. (22) recalibrated LCP-CNN (26) on the national lung screening trial (NLST) dataset to predict a 1-year cancer score, combined it with LCRAT (27), and observed that when the 66% lowest-score individuals were assigned to biennial screening, 18/195 cancers were delayed. When a maximum delay rate of 10% was assumed acceptable, the model safely extended the screening interval for 67.7% of the cases. In our study, the same threshold (90% sensitivity) resulted in a 16.6% rate of safe interval extension. In another recent study (28), researchers developed ScreenLungNet based on a commercial AI assistant, which, in a high negative predictive value (99.9%) setting, yielded a delayed cancer diagnosis rate of 2% and a specificity of 15%.

Many ethical and safety considerations arise when deploying externally validated AI algorithms in the clinical setting. Studies have reported various degrees of performance drop upon external validation of radiology AI algorithms (29,30). Despite significant improvements in the generalizability of newer algorithms, clinical deployment requires some degree of validation in that specific setting. An important decision in this process is whether to validate retrospectively or prospectively, with the former being much more feasible and the latter being more accurate (31). Another often overlooked component of investigating AI solutions is aligning research and clinical metrics (31). For instance, in our study, while Sybil demonstrated high overall AUCs, its 2-year performance in Lung-RADS 1 and 2 cases, which related most to our use case, was only modest. On the other hand, analysis of the ROC curve indicated potential clinical benefit, as a high-sensitivity, low-specificity threshold could have been beneficial. Furthermore, the interpretability limitations of deep learning algorithms warrant caution in the extent of their involvement (32). Another aspect to consider when externally validating algorithms is the effect of scan acquisition parameters. Notably, Sybil has been shown to be robust across various acquisition settings (33).

Our study has several limitations. Firstly, the data were gathered retrospectively from a single center. One consequence of this design is the use of different Lung-RADS versions, which has added heterogeneity to Lung-RADS categories. Another consequence is that not all screening scans were low-dose CTs. In addition, follow-up rates hovered around 50%, even for high-risk cases such as Lung-RADS 4a. This has most likely contributed to the small number of Lung-RADS 1 and 2 cases that developed cancer within 2 years, limiting the strength of our conclusions and analyses. While screening sessions would ideally occur annually at the same time of year, disruptions to the timeline were common. Moreover, occasionally, patients who were screened were not eligible under the USPSTF screening criteria. Finally, we could not perform a full analysis of the Brock model and had to rely on a subset of patients due to limitations in data collection.

To conclude, we demonstrated that Sybil could effectively identify very low-risk LCS subjects suitable for switching to a biennial screening schedule, effectively reducing the workload on radiology departments and reducing lung cancer mortality by freeing up space for new baseline screenings. Further research, especially in a prospective setting, is required to validate this potential. Other candidate applications of Sybil in lung cancer screening include improving risk stratification of intermediate-risk subjects (i.e., Lung-RADS 3), enhancing patient communication through providing a numerical risk score, and assisting in assigning high-risk cases to biopsy vs short-term follow-up.

## Data Availability

All data produced in the present study are available upon reasonable request to the authors

## Acknowledgments

We would like to thank Peter G. Mikhael for his contributions to this study and for providing us with the NLST data.

