## Supplementary for "Investigating AI’s potential in risk-stratifying lung cancer screenings for extending screening intervals and reducing workload: A retrospective analysis of a tertiary hospital cohort"

### Supplementary methods

**Database search:** We searched for lung cancer screening sessions using the keywords lung-rads or lungrads, or a combination of “lung cancer” and screen*, to allow for variations such as “lung cancer screening” or “screen for lung cancer”.

**Image acquisition description and statistics**

Out of 3082 scans processed by Sybil, 2889 (93.7%) were acquired on a General Electric LightSpeed VCT scanner, 129 (4.2%) on a Siemens SOMATOM Definition Flash scanner, 43 (1.4%) on a Siemens Biograph64_mCT 4R scanner, 13 (0.4%) on a Siemens SOMATOM Definition Edge scanner, 7 (0.2%) on a Siemens Biograph64 scanner, and 1 (0.0%) on a SOMATOM Definition AS+ scanner. A total of 2719 (88.2%) scans had a slice thickness of 2.5mm, 191 (6.2%) 2mm, 170 (5.5%) 1.25mm, and 2 (0.1%) 1mm.

### Supplementary results

| Table S1. Annual post-baseline lung cancer incidence | | | | | | | | | | | |
| --- | --- | --- | --- | --- | --- | --- | --- | --- | --- | --- | --- |
| Lung-RADS | New lung cancer diagnoses (incidence %, 95% CI) | | | | | | | | | | |
|  | Year 1 | Year 2 | Year 3 | Year 4 | Year 5 | Year 6 | Year 7 | Year 8 | Year 9 | Year 10 | Total (Assuming lost to follow-up as negative) |
| 0 | 2/26 (7.7%, 1.3-26.6) | 0/15 (0.0%, 0.0-25.3) | 0/12 (0.0%, 0.0-30.1) | 0/8 (0.0%, 0.0-40.2) | 0/4 (0.0%, 0.0-60.4) | 0/3 (0.0%, 0.0-69.0) | 0/2 (0.0%, 0.0-80.2) | 1/2 (50.0%, 2.7-97.3) | 0/1 (0.0%, 0.0-94.5) | 0 (n/a) | 3/32 (9.4%, 2.5-26.2) |
| 1 | 1/498 (0.2%, 0.0-1.3) | 1/379 (0.3%, 0.0-1.7) | 0/199 (0.0%, 0.0-2.4) | 1/99 (1.0%, 0.1-6.3) | 0/40 (0.0%, 0.0-10.9) | 0/23 (0.0%, 0.0-17.8) | 0/12 (0.0%, 0.0-30.1) | 0/8 (0.0%, 0.0-40.2) | 0/3 (0.0%, 0.0-69.0) | 0 (n/a) | 3/549 (0.5%, 0.1-1.7) |
| 2 | 1/1240 (0.1%, 0.0-0.5) | 8/958 (0.8%, 0.4-1.7) | 2/402 (0.5%, 0.1-2.0) | 1/181 (0.6%, 0.0-3.5) | 1/81 (1.2%, 0.1-7.6) | 0/46 (0.0%, 0.0-9.6) | 2/28 (7.1%, 1.2-25.0) | 0/18 (0.0%, 0.0-21.9) | 0/9 (0.0%, 0.0-37.1) | 0/3 (0.0%, 0.0-69.0) | 15/1363 (1.1%, 0.6-1.9) |
| 3 | 3/76 (3.9%, 1.0-11.9) | 0/49 (0.0%, 0.0-9.1) | 0/33 (0.0%, 0.0-13.0) | 1/17 (5.9%, 0.3-30.8) | 0/8 (0.0%, 0.0-40.2) | 0/5 (0.0%, 0.0-53.7) | 0/3 (0.0%, 0.0-69.0) | 0/3 (0.0%, 0.0-69.0) | 0/1 (0.0%, 0.0-94.5) | 0 (n/a) | 4/84 (4.8%, 1.5-12.4) |
| 4a | 9/52 (17.3%, 8.7-30.8) | 1/30 (3.3%, 0.2-19.1) | 0/21 (0.0%, 0.0-19.2) | 0/10 (0.0%, 0.0-34.5) | 0/7 (0.0%, 0.0-43.9) | 0/4 (0.0%, 0.0-60.4) | 0/3 (0.0%, 0.0-69.0) | 0/2 (0.0%, 0.0-80.2) | 0/1 (0.0%, 0.0-94.5) | 0 (n/a) | 10/58 (17.2%, 9.0-29.9) |
| 4b | 14/30 (46.7%, 28.8-65.4) | 0/11 (0.0%, 0.0-32.1) | 0/5 (0.0%, 0.0-53.7) | 0/2 (0.0%, 0.0-80.2) | 1/2 (50.0%, 2.7-97.3) | 0 (n/a) | 0 (n/a) | 0 (n/a) | 0 (n/a) | 0 (n/a) | 15/30 (50.0%, 31.7-68.3) |
| 4x | 5/5 (100.0%, 46.3-100.0) | 0 (n/a) | 0 (n/a) | 0 (n/a) | 0 (n/a) | 0 (n/a) | 0 (n/a) | 0 (n/a) | 0 (n/a) | 0 (n/a) | 5/9 (55.6%, 22.7-84.7) |
| Total | 35/1927 (1.8%, 1.3-2.5) | 10/1442 (0.7%, 0.4-1.3) | 2/672 (0.3%, 0.1-1.2) | 3/317 (0.9%, 0.2-3.0) | 2/142 (1.4%, 0.2-5.5) | 0/81 (0.0%, 0.0-5.6) | 2/48 (4.2%, 0.7-15.4) | 1/33 (3.0%, 0.2-17.5) | 0/15 (0.0%, 0.0-25.3) | 0/3 (0.0%, 0.0-69.0) | 55/2125 (2.6%, 2.0-3.4) |

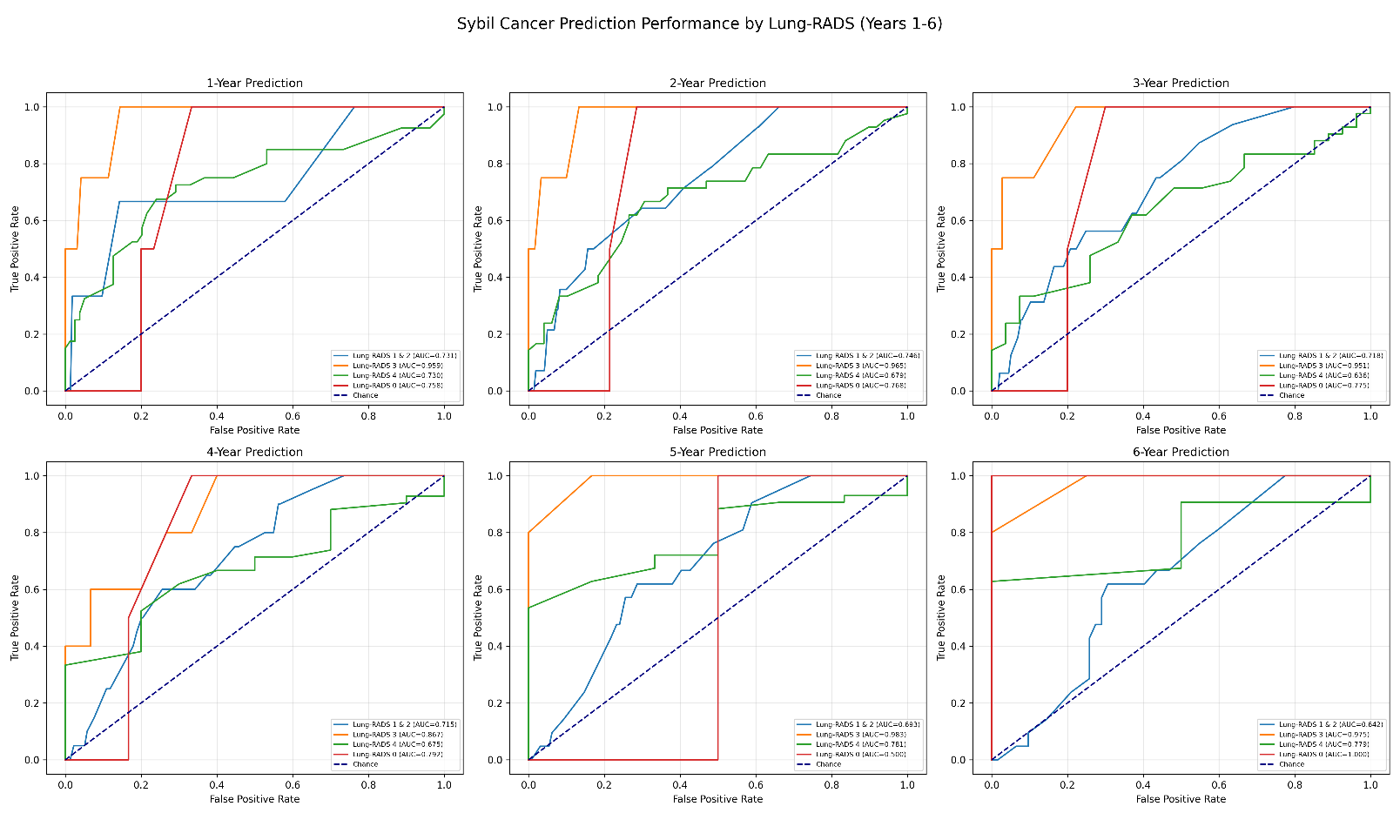

Figure S1. Sybil’s cancer performance by Lung-RADS (Years 1-6)

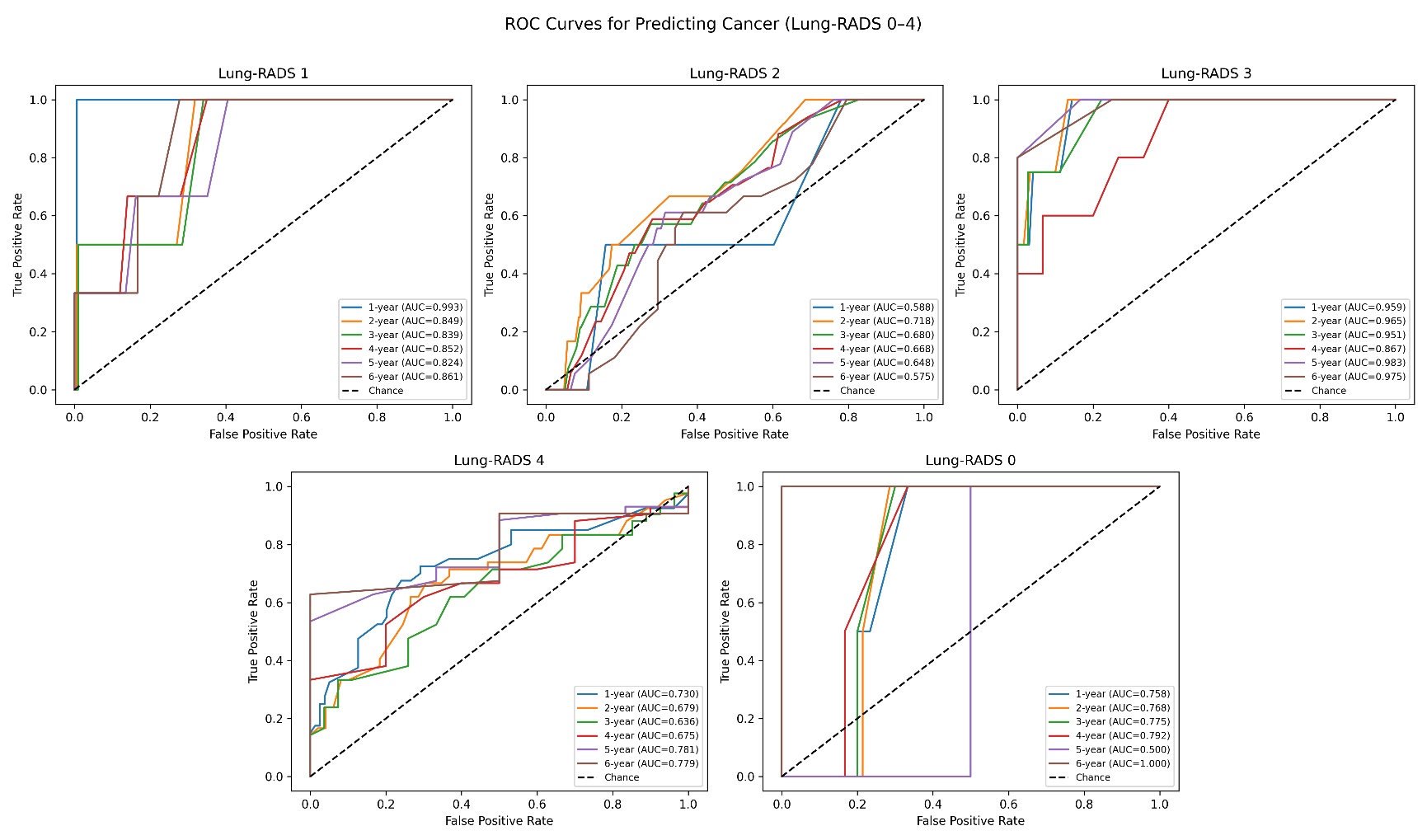

Figure S2. Sybil’s cancer performance by year (Lung-RADS 0-4)

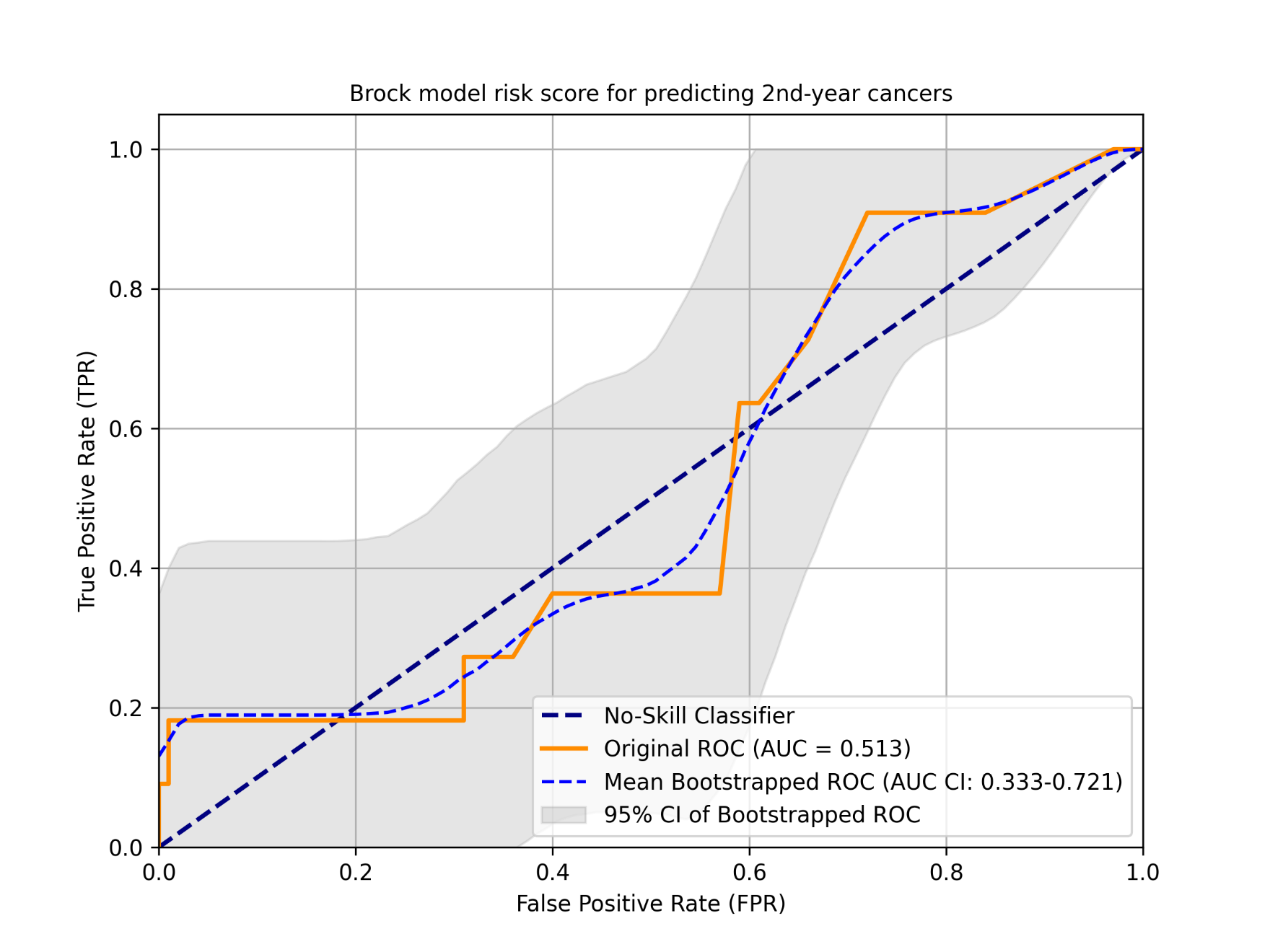

Figure S3. Brock model risk score for predicting 2nd-year cancers
